# The impact of COVID-19 on the oncologic outcomes of 3236 patients undergoing ColoRectal Cancer surgery in Northern Italy in 2019 and 2020 (COVID-CRC): results of a multicentric comparative cohort study

**DOI:** 10.1101/2021.04.19.21255730

**Authors:** Matteo Rottoli, Gianluca Pellino, Antonino Spinelli, Maria Elena Flacco, Lamberto Manzoli, Mario Morino, Salvatore Pucciarelli, Elio Jovine, Moh’d Abu Hilal, Riccardo Rosati, Alessandro Ferrero, Andrea Pietrabissa, Marcello Guaglio, Nicolò de Manzini, Pierluigi Pilati, Elisa Cassinotti, Giusto Pignata, Orlando Goletti, Enrico Opocher, Piergiorgio Danelli, Gianluca Sampietro, Stefano Olmi, Nazario Portolani, Gilberto Poggioli, the COVID-CRC Collaborative

**Author notes:** Corresponding author (and requests for reprints): Matteo Rottoli, Surgery of the Alimentary Tract, IRCCS Azienda Ospedaliero Universitaria di Bologna, Department of Medical and Surgical Science, Alma Mater Studiorum University of Bologna, Via Massarenti 9, 40138 Bologna, Italy. Collaborating authors are listed in the Supplementary File 2.

## Abstract

**Objective:** This study compared all patients undergoing surgery for colorectal cancer in 20 hospitals of Northern Italy in 2019 versus 2020, in order to evaluate whether COVID-19-related delays in the execution of colorectal cancer screening resulted in more advanced cancers at diagnosis and worse clinical outcomes.

**Design:** A retrospective multicentric cohort analysis of patients who underwent surgery for colorectal cancer in March-December 2019 (2019) versus March-December 2020 (2020). The independent predictors of disease stage (oncologic stage, associated symptoms, clinical T4 stage, metastasis) and postoperative outcome (surgical complications, palliative surgery, 30-day death) were evaluated using logistic regression.

**Results:** The sample consisted of 1755 patients operated in 2019, and 1481 in 2020 (both mean ages 69.6 years). The proportions of cancers with symptoms, clinical T4 stage, liver and lung metastases in 2019 and 2020 were, respectively: 80.8% vs 84.5%; 6.2% vs 8.7%; 10.2% vs 10.3%; and 3.0% vs 4.4%. The proportions of surgical complications, palliative surgery, and death in 2019 and 2020 were, respectively: 34.4%vs 31.9%; 5.0% vs 7.5%; and 1.7% vs 2.4%. At multivariate analysis, as compared with 2019, cancers in 2020 were significantly more likely to be symptomatic (Odds Ratio - OR: 1.36, 95% Confidence Interval - CI: 1.09-1.69), in clinical T4 stage (OR: 1.38; 1.03-1.85), with multiple liver metastases (OR: 2.21; 1.24-3.94), but less likely to cause surgical complications (OR: 0.79; 0.68-0.93).

**Conclusions:** Colorectal cancer patients who had surgery between March and December 2020 had an increased risk of more advanced disease in terms of associated symptoms, cancer location, clinical T4 stage, and number of liver metastases.

**SHORT SUMMARY BOX:** 

**What is already known about this subject?:** A specific search regarding the correlation between colorectal cancer oncologic outcomes and COVID-19 showed a few modeling studies which reported the predictions of the potential impact of the diagnostic delays (due to the reduction of the screening programs) on the survival of patients affected by colorectal cancer. However, no study reported any real-life evidence regarding the correlation between the COVID-19 outbreak and the deteriorations of the oncologic outcomes of patients with colorectal cancer.

**What are the new findings?:** The present study showed that patients who had surgery for colorectal cancer between March and December 2020 had an increased risk of more advanced disease in terms of associated symptoms, cancer location, clinical T4 stage, and number of liver metastases, than patients who had surgery between March and December 2019.

**How might it impact on clinical practice in the foreseeable future?:** The present study confirmed that the backlogs of the screening programs have had, and probably will have, detrimental effects on the oncologic outcomes of patients affected by colorectal cancer. Increased resources should be placed in order to reactivate and enhance the screening programs, and to reduce the risk of colorectal cancer patients to be diagnosed with advanced cancer in the next future.

## INTRODUCTION

Coronavirus disease 19 (COVID-19), which is associated with severe acute respiratory syndrome coronavirus 2 (SARS-CoV-2) infection, has spread worldwide since it was first reported in China in December 2019.^1,2^ Italy has witnessed a rapid and uncontrolled spread of the infection since February 2020, and a number of related deaths which surpassed those of China by the end of March 2020, especially in the northern regions.^3,4^ Due to the great pressure on the healthcare system in terms of the increased amount of resources required for the diagnosis and the treatment of COVID-19 patients, a national lockdown was established on March 10, 2020.^5^ As a consequence, elective surgical activities were greatly reduced, and screening programs were suspended for the greater part of the period between March and May 2020.^6^ This included the faecal immunochemical test (FIT) which has been widely adopted in Italy and many European countries as screening for colorectal lesions.^7^ A similar scenario has been observed in other countries,^8,9^ and has raised concerns regarding the risk of delayed diagnoses of precancerous lesions and early cancers^10^ and, therefore, of more advanced stages of colorectal cancer at surgery which could influence a significant increase in mortality.^11–13^

However, to date, no evidence has been provided regarding an increase in the advanced oncologic stage in patients who underwent surgery for colorectal cancer in 2020.

The aim of the present study was therefore to analyse the outcomes of patients undergoing surgery for colorectal cancer in Northern Italy between March and December 2020, and to compare them to those of patients with the same diagnosis who had had surgery in the same period of 2019.

## METHODS

### Study design and participants

This is a retrospective cohort study enrolling all adult (≥ 18 years) patients who underwent surgery for a proven or suspected colorectal malignancy, and had been followed for at least 30 days postoperatively, from 1 March to 31 December 2019 and from 1 March to 31 December 2020, in 20 referral centres for the treatment of colorectal cancer located in the Italian regions of Lombardy, Piedmont, Emilia-Romagna, Veneto and Friuli-Venezia-Giulia. The details of the centres are listed in Supplementary Table 1.

The study was approved by the Ethical Committee of the leading centre (Azienda Ospedaliero Universitaria di Bologna, Alma Mater Studiorum University of Bologna) and carried out in the participating centres according to local regulations. The study was registered on ClinicalTrials.gov (COVID-CRC, NCT04712292).

The inclusion criteria were: a preoperative or postoperative histologically confirmed diagnosis of cancer; elective or urgent surgery; palliative or curative surgery; location of the cancer in the colon, the rectum or the anus and any type of surgery, including surgical exploration or palliative procedures. The exclusion criteria were: recurrent colorectal cancer after previous surgery; cancer originating from other organs; lack of significant histological details (except when the cancer was not removed, i.e. the palliative procedures, carcinosis, etc.) and lack of a 30-day follow-up.

All patients were included in the study regardless of the 30-day outcome (discharge, still in the hospital or death) and all data were extracted directly from the charts, validated by trained specialist physicians in the participating centres, and entered in REDCap software (Research Electronic Data Capture).^14^

The dataset included details regarding patient history, comorbidities, preoperative diagnosis, the use of neoadjuvant therapy, surgical procedures, the onset of 30-day postoperative complications and mortality, location and histological examination. The biology of the tumour was considered aggressive if any of the following characteristics were found at histological examination: the presence of signet ring cells, a mucinous tumour, tumour budding, lymphovascular invasion, perineural invasion, and lymphangitis. Concerning location, right colon cancers included those in the cecum, the ascending and the transverse colon proximally to the splenic flexure; left colon included those located between the splenic flexure and the rectosigmoid junction; and rectum cancers included those located distally to the rectosigmoid junction, including the anus.

The study outcomes included the following measures of cancer clinical stage or outcome:

1. associated symptoms at diagnosis (including bleeding, change in bowel habits, tenesmus, anaemia, abdominal pain, weight loss, bowel obstruction);
2. clinical T4 stage (defined as the presence of cancer-induced spiculations extended over the bowel wall or suspicion of infiltration of the surrounding organs or structures at preoperative radiological examination);
3. advanced TNM stage (cancers with T4N0, any T N1 or N2, any T any N M+ stages; plus all cases without final histology which required palliative surgery);
4. presence of lung metastases;
5. presence of liver metastases (and, among these cases, the proportion of patients who were diagnosed with more than one metastasis);
6. surgical complications;
7. palliative surgery (defined as any procedure which did not have the aim of radically removing the primary cancer, either planned preoperatively in order to reduce the associated symptoms or to confirm the diagnosis, or which became necessary due to unexpected findings during surgery; the presence of distant metastases did not define palliative surgery per se as long as the surgical procedure was carried out according to the oncologic principles of radical surgery);
8. emergency surgery, including all cases which required surgery within 48 hours from the admission to hospital;
9. 30-day death.

### Statistical analysis

The continuous variables were expressed as mean ± standard deviation (SD) while the categorical variables were presented as number (%).

For each of the nine outcomes, the differences in categorical and continuous variables between 2020 and 2019 were initially evaluated using the chi-squared test and the t-test, respectively. The potential independent predictors of outcomes 1-3 5-7 and 9 were then evaluated using logistic regression while no multivariable analysis was attempted to predict lung metastases and emergency surgery, due to the scarce number of positive/negative observations (3.7% and 9.8% of the sample, respectively).

To reduce overfitting, all the models were built including only the variables which were significant at the 0.1 level at the univariate analyses, with the exception of age, gender, year (2020 versus 2019), region (Lombardy versus others) and cancer site (rectum versus others) which were included a priori. Given that the clinical T4 stage, advanced cancer and liver metastases were highly collinear, three separate models were fit for each outcome, each including only one of the three covariates. The model with the highest pseudo-R2 was kept as final. In addition, all the models were repeated with the same covariates, including region as a cluster variable,^15^ with no substantial changes in the final estimates; they were thus not shown to avoid redundancy.

Standard diagnostic procedures were adopted to check the validity of all the models: influential observation analysis (Dbeta, change in Pearson chi-square), the Hosmer-Lemeshow test for the goodness of fit and C statistic (area under the Receiving Operator Curve). Missing data were <5% in all the primary analyses; thus, no missing imputation technique was adopted. Statistical significance was defined as a two-sided p-value<0.05; all the analyses were carried out using Stata, version 13.1 (Stata Corp., College Station, TX, 2014).

### Patient and public involvement statement

Patients and public were not involved in the development of the present study.

## RESULTS

After the exclusion of 52 patients who met the exclusion criteria, 3236 cases were analysed. Of these, 1755 (54.2%) had undergone surgery between March and December 2019 while 1481 (45.8%) had received their treatment between March and December 2020. The proportion of patients who had undergone surgery during the lockdown period (1 March - 31 May) in 2020 was similar to that of the previous year (32.7% vs. 32.5%), suggesting that the treatment of patients affected by colorectal cancer was not postponed, despite the fact that elective activity was severely reduced.

As shown in Table 1, the proportion of patients treated in each region in 2020 was comparable to that of 2019, except for the hospitals located in Lombardy (55.6% vs. 49.4%, p=0.011). The details of each centre in terms of variation of the volume of patients is shown in Supplementary Table 1.

**Table 1.** Characteristics and outcomes of the sample, overall and by year of surgical procedure (2020 vs 2019). Abbreviations: BMI: body mass index; COPD: chronic obstructive pulmonary disease; ASA: American Society of Anesthesiologists * Chi-squared test for categorical variables, t-test for continuous variables. ** Including only the 331 patients with liver metastasis.

|  | **Total**<br>**sample**<br>**(n=3236)** | **March-December**<br>**2019**<br>**(n=1755)** | **March-December**<br>**2020**<br>**(n=1481)** | **p** |
| --- | --- | --- | --- | --- |
| Mean age in years (SD) | 69.6 (13.0) | 69.6 (12.8) | 69.6 (13.2) | 0.9 |
| Male gender, % | 42.9 | 42.9 | 42.9 | 0.9 |
| Mean BMI in kg/m<sup>2</sup> (SD) | (n=2854)<br>25.3 (4.9) | (n=1558)<br>25.3 (4.8) | (n=1296)<br>25.3 (5.0) | 0.9 |
| Smoking status, % | (n=2743) | (n=1521) | (n=1222) |  |
| - Never | 60.7 | 60.0 | 61.5 | 0.6 |
| - Past | 25.6 | 25.1 | 26.4 | 0.7 |
| - Current | 13.7 | 14.9 | 12.1 | 0.4 |
| Family history of cancer, % | (n=2793)<br>12.8 | (n=1508)<br>13.1 | (n=1285)<br>12.5 | 0.6 |
| Comorbidities, % |  |  |  |  |
| - Myocardial infarction | 54.2 | 52.1 | 56.8 | 0.007 |
| - Type II Diabetes | 15.9 | 16.6 | 15.0 | 0.2 |
| - COPD | 10.3 | 10.3 | 10.4 | 0.9 |
| - Stroke | 6.3 | 5.9 | 6.9 | 0.2 |
| - Renal disease | 5.2 | 4.7 | 5.7 | 0.2 |
| - Other malignancies | 11.0 | 10.7 | 11.3 | 0.5 |
| - Other colorectal cancer | 3.2 | 3.2 | 3.3 | 0.9 |
| Primary rectal cancer site, % | 30.8 | 28.3 | 33.8 | 0.001 |
| Neoadjuvant therapy in rectal cancer, % | (n=1016)<br>52.1 | (n=503)<br>51.9 | (n=513)<br>52.2 | 0.9 |
| ASA score >2, % | 44.4 | 42.4 | 46.7 | 0.015 |
| Aggressive tumour biology, % | 73.0 | 71.9 | 74.4 | 0.10 |
| Hospital site (Region), % |  |  |  |  |
| - Lombardy | 52.8 | 55.6 | 49.4 | 0.011 |
| - Emilia-Romagna | 15.8 | 15.2 | 16.6 | 0.7 |
| - Piedmont | 15.2 | 14.8 | 15.5 | 0.9 |
| - Veneto | 12.4 | 10.7 | 14.3 | 0.3 |
| - Friuli-Venezia-Giulia | 3.8 | 3.7 | 4.2 | 0.9 |
| Faecal blood test carried out, % |  |  |  |  |
| - Overall | 25.5 | 26.6 | 24.3 | 0.13 |
| - Among asymptomatic subjects only | 12.7 | 14.4 | 10.7 | 0.002 |
| *Outcomes:* |  |  |  |  |
| Symptoms at diagnosis, % | 82.5 | 80.8 | 84.5 | 0.006 |
| Clinical T4 stage, % | 7.4 | 6.2 | 8.7 | 0.008 |
| Cancer stage, % |  |  |  | 0.6 |
| - Early | 51.4 | 51.7 | 50.9 |  |
| - Advanced | 48.6 | 48.3 | 49.1 |  |
| Liver metastasis, % | (n=3101)<br>10.2 | (n=1642)<br>10.2 | (n=1459)<br>10.3 | 0.9 |
| Multiple liver metastases, % ** | (n=331)<br>76.7 | (n=179)<br>72.1 | (n=152)<br>82.2 | 0.029 |
| Lung metastasis, % | 3.7 | 3.0 | 4.4 | 0.038 |
| Surgical complications, % | 33.3 | 34.4 | 31.9 | 0.15 |
| Palliative surgery, % | 6.2 | 5.0 | 7.5 | 0.003 |
| Emergency surgery, % | 90.2 | 91.0 | 89.2 | 0.08 |
| 30-day death, % | 2.0 | 1.7 | 2.4 | 0.15 |

In 2020, a greater proportion of patients diagnosed with rectal cancer (33.8% vs. 28.3%, p=0.001) and a greater proportion of patients with an ASA (American Society of Anesthesiologists) score higher that 2 (46.7% vs. 42.4%, p=0.015) were observed. While the rates of early/advanced cancers were comparable between the two years (the classification of the cases according to the American Joint Committee on Cancer - AJCC - stages is shown in Table 2), a significantly higher rate of clinical T4 cancers was found in 2020 (8.7% vs. 6.2%, p=0.008).

**Table 2.**
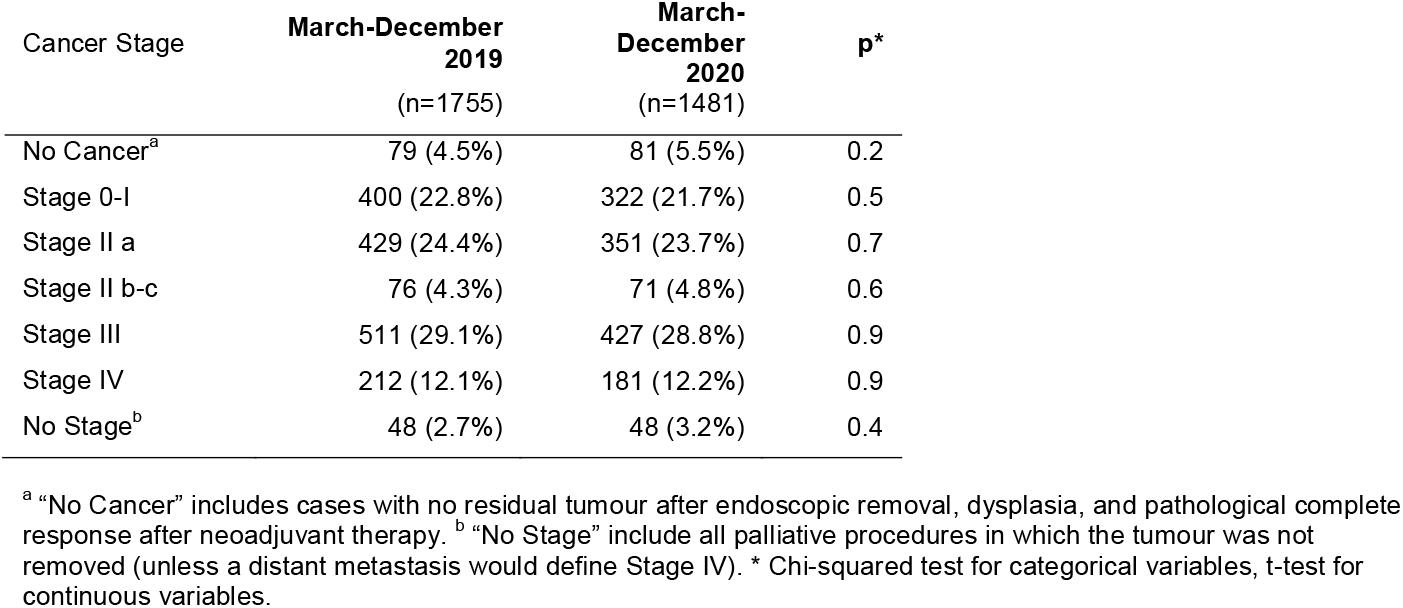
Distribution of the oncologic stages according to the American Joint Committee on Cancer (AJCC).

| Cancer Stage | March-December<br>2019<br>(n=1755) | March-December<br>2020<br>(n=1481) | p* |
| --- | --- | --- | --- |
| No Cancer<sup>a</sup> | 79 (4.5%) | 81 (5.5%) | 0.2 |
| Stage 0-I | 400 (22.8%) | 322 (21.7%) | 0.5 |
| Stage II a | 429 (24.4%) | 351 (23.7%) | 0.7 |
| Stage II b-c | 76 (4.3%) | 71 (4.8%) | 0.6 |
| Stage III | 511 (29.1%) | 427 (28.8%) | 0.9 |
| Stage IV | 212 (12.1%) | 181 (12.2%) | 0.9 |
| No Stage<sup>b</sup> | 48 (2.7%) | 48 (3.2%) | 0.4 |

<sup>a</sup> "No Cancer" includes cases with no residual tumour after endoscopic removal, dysplasia, and pathological complete response after neoadjuvant therapy. <sup>b</sup> "No Stage" include all palliative procedures in which the tumour was not removed (unless a distant metastasis would define Stage IV). \* Chi-squared test for categorical variables, t-test for continuous variables.

The proportion of patients diagnosed with liver metastases (10.3% vs 10.2%, p= 0.9) was similar in 2020 and 2019 while a greater rate of lung metastases was diagnosed in 2020 (4.4% vs. 3.3%, p=0.038). A significantly higher rate of patients were diagnosed with more than one liver metastasis in 2020 than in 2019 (82.2% vs. 72.1%, p=0.029) In 2020, a significant decrease in the number of patients who were diagnosed with colorectal cancer by means of screening programs without any correlated symptoms (15.5% vs. 19.9%, p=0.006) was observed while a greater proportion of the surgical procedures were carried out with a palliative intent (7.5% vs. 5%, p=0.003).

At the multivariate analysis assessing the potential predictors of the oncologic (Table 3A) and the perioperative (Table 3B) outcomes, undergoing surgery in 2020 was significantly associated with a higher rate of symptomatic cancers (OR 1.36, 95% CI 1.09-1.69, p=0.006), a higher proportion of clinical T4 stage tumours (OR 1.40, 95% CI 1.04-1.87, p=0.024), and a lower risk of postoperative surgical complications (OR 0.79, 95% CI 0.68-0.93, p=0.004). Among all the variables, a clinical T4 stage was significantly associated with 30-day mortality (OR 3.87, 95% CI 2.06-7.28, p<0.001), postoperative complications (OR 2.02, 95% CI 1.51-2.70, p<0.001), palliative surgery (OR 8.22, 95% CI 5.65-11.9, p<0.001) and liver metastases (OR 2.47, 95% CI 1.73-3.52, p<0.001).

**Table 3A.** Multivariate analyses evaluating the potential predictors of each oncologic outcome at presentation.

| Variables | Symptoms<br>at diagnosis<br>(n=2671) |  |  | Clinical<br>T4 stage<br>(n=229) |  |  | Advanced<br>stage<br>(n=1574) |  |  | Liver<br>metastasis<br>(n=331) |  |  |
| --- | --- | --- | --- | --- | --- | --- | --- | --- | --- | --- | --- | --- |
|  | % | OR (95% CI) | p<sup>A</sup> | % | OR (95% CI) | p<sup>A</sup> | % | OR (95% CI) | p<sup>A</sup> | % | OR (95% CI) | p<sup>A</sup> |
| Year, % |  |  |  |  |  |  |  |  |  |  |  |  |
| - 2019 | 80.8 | 1 (ref. cat.) | -- | 6.2 | 1 (ref. cat.) | -- | 48.3 | 1 (ref. cat.) | -- | 10.2 | 1 (ref. cat.) | -- |
| - 2020 | 84.5 | 1.36 (1.09-1.69) | 0.006 | 8.7 | 1.38 (1.03-1.85) | 0.029 | 49.1 | 1.01 (0.86-1.18) | 0.9 | 10.3 | 0.98 (0.77-1.25) | 0.9 |
| Age class in years |  |  |  |  |  |  |  |  |  |  |  |  |
| - <60 | 81.1 | 1 (ref. cat.) | -- | 7.7 | 1 (ref. cat.) | -- | 52.7 | 1 (ref. cat.) | -- | 14.0 | 1 (ref. cat.) | -- |
| - 60-69.9 | 72.6 | 0.59 (0.44-0.79) | 0.001 | 7.6 | 1.06 (0.69-1.63) | 0.8 | 48.1 | 0.91 (0.72-1.16) | 0.4 | 11.6 | 0.81 (0.57-1.14) | 0.2 |
| - 70-79.9 | 83.6 | 1.07 (0.78-1.48) | 0.7 | 7.2 | 0.93 (0.61-1.41) | 0.7 | 48.2 | 0.75 (0.60-0.94) | 0.013 | 9.3 | 0.56 (0.40-0.79) | 0.001 |
| - ≥80 | 91.1 | 2.11 (1.42-3.11) | <0.001 | 7.2 | 0.91 (0.57-0.44) | 0.7 | 46.1 | 0.58 (0.45-0.75) | <0.001 | 10.8 | 0.35 (0.23-0.52) | <0.001 |
| Age, 10-year increase | -- | 1.01 (1.00-1.02) | 0.004 | -- | 0.99 (0.98-1.01) | 0.3 | -- | 0.98 (0.97-0.99) | <0.001 | -- |  |  |
| Gender |  |  |  |  |  |  |  |  |  |  |  |  |
| - Female | 81.8 | 1 (ref. cat.) | -- | 6.7 | 1.33 (0.99-1.78) | 0.056 | 47.8 | 1 (ref. cat.) | -- | 10.9 | 1 (ref. cat.) | -- |
| - Male | 83.6 | 1.28 (1.03-1.59) | 0.027 | 8.3 |  |  | 49.7 | 1.04 (0.89-1.23) | 0.6 | 9.4 | 0.85 (0.66-1.10) | 0.2 |
| Current smoker, % |  |  |  |  |  |  |  |  |  |  |  |  |
| - No | 83.1 | -- | -- | 7.5 | -- | -- | 48.2 |  |  | 10.3 | -- | -- |
| - Yes | 82.1 | -- | -- | 7.3 | -- | -- | 45.7 |  |  | 9.6 | -- | -- |
| Family history of cancer |  |  |  |  |  |  |  |  |  |  |  |  |
| - No | 82.5 | 1 (ref. cat.) | -- | 7.8 | -- | -- | 49.2 | 1 (ref. cat.) | -- | 10.3 | -- | -- |
| - Yes | 76.3 | 0.65 (0.48-0.87) | 0.004 | 4.9 | -- | -- | 42.7 | 0.73 (0.58-0.93) | 0.011 | 10.3 | -- | -- |
| Diabetes |  |  |  |  |  |  |  |  |  |  |  |  |
| - No | 81.9 | 1 (ref. cat.) | -- | 7.4 | -- | -- | 48.8 | -- | -- | 10.4 | -- | -- |
| - Yes | 85.8 | 1.36 (0.98-1.89) | 0.07 | 7.1 | -- | -- | 47.7 | -- | -- | 9.5 | -- | -- |
| Myocardial infarction |  |  |  |  |  |  |  |  |  |  |  |  |
| - No | 79.3 | 1 (ref. cat.) | -- | 8.1 | -- | -- | 49.4 | -- | -- | 12.0 | 1 (ref. cat.) | -- |
| - Yes | 85.2 | 1.25 (0.98-1.59) | 0.07 | 6.8 | -- | -- | 48.0 | -- | -- | 8.7 | 0.80 (0.60-1.06) | 0.12 |
| Stroke |  |  |  |  |  |  |  |  |  |  |  |  |
| - No | 82.1 | 1 (ref. cat.) | -- | 7.6 | -- | -- | 49.0 | -- | -- | 10.5 | -- | -- |
| - Yes | 88.8 | 1.58 (0.92-2.73) | 0.09 | 5.0 | -- | -- | 43.9 | -- | -- | 6.8 | -- | -- |
| Other cancers |  |  |  |  |  |  |  |  |  |  |  |  |
| - No | 83.3 | 1 (ref. cat.) | -- | 7.3 | -- | -- | 49.1 | -- | -- | 10.4 | -- | -- |
| - Yes | 76.6 | 0.46 (0.34-0.63) | <0.001 | 8.0 | -- | -- | 45.1 | -- | -- | 9.0 | -- | -- |
| Hospital in Lombardy, % |  |  |  |  |  |  |  |  |  |  |  |  |
| - No | 79.2 | 1 (ref. cat.) | -- | 8.9 | 1 (ref. cat.) | -- | 47.1 | 1 (ref. cat.) | -- | 11.1 | 1 (ref. cat.) | -- |
| - Yes | 85.5 | 1.36 (1.09-1.69) | 0.001 | 6.0 | 0.62 (0.46-0.83) | 0.002 | 50.0 | 1.18 (1.01-1.38) | 0.046 | 9.4 | 0.87 (0.68-1.12) | 0.3 |
| Fecal blood test, % |  |  |  |  |  |  |  |  |  |  |  |  |
| -No | 88.6 | 1 (ref. cat.) | -- | 8.5 | 1 (ref. cat.) | -- | 51.3 | 1 (ref. cat.) | -- | 11.5 | 1 (ref. cat.) | -- |
| -Yes | 64.5 | 0.26 (0.21-0.33) | <0.001 | 3.8 | 0.50 (0.33-0.76) | 0.002 | 39.8 | 0.66 (0.54-0.79) | <0.001 | 6.4 | 0.60 (0.43-0.83) | 0.002 |
| Rectum location, % |  |  |  |  |  |  |  |  |  |  |  |  |
| - No | 80.9 | 1 (ref. cat.) | -- | 7.3 | 1 (ref. cat.) | -- | 50.0 | 1 (ref. cat.) | -- | 9.9 | 1 (ref. cat.) | -- |
| - Yes | 85.8 | 1.92 (1.49-2.47) | <0.001 | 7.4 | 1.10 (0.80-1.51) | 0.6 | 44.8 | 0.87 (0.73-1.05) | 0.14 | 10.6 | 1.03 (0.79-1.35) | 0.8 |
| ASA score >2, % |  |  |  |  |  |  |  |  |  |  |  |  |
| - No | 79.3 | 1 (ref. cat.) | -- | 6.4 | 1 (ref. cat.) | -- | 46.7 | 1 (ref. cat.) | -- | 9.1 | 1 (ref. cat.) | -- |
| - Yes | 86.6 | 1.05 (0.87-1.27) | 0.5 | 8.6 | 1.11 (0.80-1.51) | 0.6 | 51.1 | 1.26 (1.06-1.50) | 0.009 | 11.6 | 1.62 (1.23-2.14) | 0.001 |
| Aggressive cancer biology, % |  |  |  |  |  |  |  |  |  |  |  |  |
| - No | 75.1 | 1 (ref. cat.) | -- | 4.4 | 1 (ref. cat.) | -- | 28.1 | 1 (ref. cat.) | -- | 6.4 | 1 (ref. cat.) | -- |
| - Yes | 85.3 | 2.04 (1.62-2.56) | <0.001 | 8.5 | 1.10 (0.73-1.66) | 0.6 | 56.2 | 3.65 (3.01-4.42) | <0.001 | 11.6 | 2.06 (1.47-2.87) | <0.001 |
| Clinical T4 stage, %<sup>B</sup> |  |  |  |  |  |  |  |  |  |  |  |  |
| - No | 81.3 | 1 (ref. cat.) | -- | -- | -- | -- | 45.1 | 1 (ref. cat.) | -- | 9.0 | 1 (ref. cat.) | -- |
| - Yes | 92.1 | 1.97 (1.15-3.36) | 0.013 | -- | -- | -- | 87.3 | 7.41 (4.79-11.5) | <0.001 | 21.8 | 2.47 (1.73-3.52) | <0.001 |
| Advanced stage, %<sup>B</sup> |  |  |  |  |  |  |  |  |  |  |  |  |
| - No | 78.0 | -- | -- | 1.8 | 1 (ref. cat.) | -- | -- | -- | -- | 1.0 | -- | -- |
| - Yes | 87.3 | -- | -- | 13.4 | 7.33 (4.85-11.0) | <0.001 | -- | -- | -- | 20.0 | -- | -- |
| Liver metastasis, %<sup>B</sup> |  |  |  |  |  |  |  |  |  |  |  |  |
| - No | 81.7 | -- | -- | 6.4 | -- | -- | 43.3 | -- | -- | -- | -- | -- |
| - Yes | 90.0 | -- | -- | 16.2 | -- | -- | 95.2 | -- | -- | -- | -- | -- |
| Surgical complications, % |  |  |  |  |  |  |  |  |  |  |  |  |
| - No | 81.5 | 1 (ref. cat.) | -- | 5.5 | 1 (ref. cat.) | -- | 46.6 | 1 (ref. cat.) | -- | 9.3 | 1 (ref. cat.) | -- |
| - Yes | 86.4 | 0.92 (0.73-1.15) | 0.4 | 11.2 | 1.93 (1.44-2.59) | <0.001 | 52.7 | 1.22 (1.03-1.44) | 0.023 | 12.1 | 1.21 (0.94-3.56) | 0.14 |

Abbreviations: OR: Odds Ratio; CI: Confidence Interval; ASA: American Society of Anesthesiologists. In all models, age, gender, year, region (Lombardy versus others) and cancer site (rectum versus other sites) were included a priori. <sup>A</sup> No variable other than those significant at univariate analysis was included in the final model. <sup>B</sup> Given the multicollinearity across T4 stage, advanced stage and liver metastases, three separate models were fit, each including only one of the three covariates. The model with the highest R<sup>2</sup> was kept as final. All the models were repeated with the same covariates, including region as a cluster variable, with no substantial changes in the final estimates. They were thus not shown to avoid redundancy. In all the univariate analyses, significant results (p<0.05) are indicated in bold.

**Table 3B.** Multivariate analyses evaluating the potential predictors of each recorded perioperative outcome.

| Variables | Surgical<br>Complications<br>(n=1076) |  |  | Palliative<br>Surgery<br>(n=199) |  |  | 30-day death<br>(n=64) |  |  |
| --- | --- | --- | --- | --- | --- | --- | --- | --- | --- |
|  | % | OR (95% CI) | p<sup>A</sup> | % | OR (95% CI) | p<sup>A</sup> | % | OR (95% CI) | p<sup>A</sup> |
| Year, % |  |  |  |  |  |  |  |  |  |
| - 2019 | 34.4 | 1 (ref. cat.) | -- | **5.0** | 1 (ref. cat.) | -- | 1.7 | 1 (ref. cat.) | -- |
| - 2020 | 31.9 | 0.79 (0.68-0.93) | 0.004 | **7.5** | 1.34 (0.96-1.88) | 0.08 | 2.4 | 1.26 (0.74-2.12) | 0.4 |
| Age class in years |  |  |  |  |  |  |  |  |  |
| - <60 | **29.9** | 1 (ref. cat.) | -- | 5.8 | 1 (ref. cat.) | -- | 0.1 | -- | -- |
| - 60-69.9 | **30.7** | 0.92 (0.72-1.18) | 0.7 | 5.9 | 1.03 (0.61-2.73) | 0.9 | 0.6 | -- | -- |
| - 70-79.9 | **34.6** | 1.02 (0.80-1.29) | 0.6 | 5.3 | 0.68 (0.41-1.14) | 0.15 | 2.4 | -- | -- |
| - ≥80 | **36.7** | 0.97 (0.74-1.26) | 0.8 | 7.8 | 0.89 (0.53-1.50) | 0.7 | 4.3 | -- | -- |
| Age, 10-year increase | -- | 1.01 (0.94-1.09) | 0.7 | -- | 1.08 (0.92-1.27) | 0.3 | -- | 1.87 (1.37-2.56) | <0.001 |
| Gender |  |  |  |  |  |  |  |  |  |
| - Female | **35.8** | 1 (ref. cat.) | -- | 6.2 | 1 (ref. cat.) | -- | 2.2 | 1 (ref. cat.) | -- |
| - Male | **29.8** | 0.78 (0.67-0.92) | 0.003 | 6.1 | 0.91 (0.65-1.28) | 0.08 | 1.7 | 0.79 (0.46-1.35) | 0.4 |
| Current smoker, % |  |  |  |  |  |  |  |  |  |
| - No | 33.6 | -- | -- | 5.7 | -- | -- | 2.1 | -- | -- |
| - Yes | 33.1 | -- | -- | 5.9 | -- | -- | 1.9 | -- | -- |
| Family history of cancer |  |  |  |  |  |  |  |  |  |
| - No | 34.0 | -- | -- | 6.1 | -- | -- | **2.1** | -- | -- |
| - Yes | 30.7 | -- | -- | 5.3 | -- | -- | **0.6** | -- | -- |
| Diabetes |  |  |  |  |  |  |  |  |  |
| - No | **32.3** | 1 (ref. cat.) | -- | 6.2 | -- | -- | 1.8 | -- | -- |
| - Yes | **38.1** | 1.00 (0.81-1.24) | 0.9 | 6.0 | -- | -- | 2.7 | -- | -- |
| Myocardial infarction |  |  |  |  |  |  |  |  |  |
| - No | **29.4** | 1 (ref. cat.) | -- | 5.9 | -- | -- | **0.7** | -- | -- |
| - Yes | **36.5** | 1.17 (0.98-1.40) | 0.2 | 6.4 | -- | -- | **3.0** | -- | -- |
| Stroke |  |  |  |  |  |  |  |  |  |
| - No | **32.7** | 1 (ref. cat.) | -- | 6.1 | -- | -- | **1.8** | -- | -- |
| - Yes | **41.5** | 1.25 (0.91-1.71) | 0.2 | 7.3 | -- | -- | **4.9** | -- | -- |
| Other cancers |  |  |  |  |  |  |  |  |  |
| - No | 32.7 | -- | -- | 6.2 | -- | -- | 1.8 | -- | -- |
| - Yes | 37.8 | -- | -- | 5.9 | -- | -- | 3.1 | -- | -- |
| Hospital in Lombardy, % |  |  |  |  |  |  |  |  |  |
| - No | **38.0** | 1 (ref. cat.) | -- | 5.4 | 1 (ref. cat.) | -- | 2.4 | 1 (ref. cat.) | -- |
| - Yes | **29.0** | 0.73 (0.62-0.85) | <0.001 | 6.9 | 1.55 (1.09-2.18) | 0.013 | 1.6 | 0.89 (0.53-1.53) | 0.7 |
| Fecal blood test, % |  |  |  |  |  |  |  |  |  |
| -No | **35.0** | 1 (ref. cat.) | -- | **6.8** | 1 (ref. cat.) | -- | 2.2 | -- | -- |
| -Yes | **28.8** | 0.88 (0.73-1.06) | 0.2 | **3.0** | 0.57 (0.35-0.92) | 0.019 | 1.2 | -- | -- |
| Rectum location, % |  |  |  |  |  |  |  |  |  |
| - No | 32.8 | 1 (ref. cat.) | -- | 6.0 | 1 (ref. cat.) | -- | 2.1 | 1 (ref. cat.) | -- |
| - Yes | 34.9 | 1.21 (1.01-1.44) | 0.034 | 4.8 | 0.76 (0.52-1.12) | 0.17 | 1.8 | 1.29 (0.71-2.34) | 0.4 |
| ASA score >2, % |  |  |  |  |  |  |  |  |  |
| - No | **26.5** | 1 (ref. cat.) | -- | **3.8** | 1 (ref. cat.) | -- | **0.3** | 1 (ref. cat.) | -- |
| - Yes | **41.9** | 1.74 (1.45-2.08) | <0.001 | **9.2** | 2.40 (1.64-3.52) | <0.001 | **4.1** | 8.72 (3.39-22.5) | 0.001 |
| Aggressive cancer biology, % |  |  |  |  |  |  |  |  |  |
| - No | **29.0** | 1 (ref. cat.) | -- | 6.3 | -- | -- | **1.2** | -- | -- |
| - Yes | **34.8** | 1.16 (0.95-1.40) | 0.14 | 6.1 | -- | -- | **2.3** | -- | -- |
| Clinical T4 stage, % <sup>B</sup> |  |  |  |  |  |  |  |  |  |
| - No | **32.1** | 1 (ref. cat.) | -- | **4.3** | 1 (ref. cat.) | -- | **1.6** | 1 (ref. cat.) | -- |
| - Yes | **50.7** | 2.02 (1.51-2.70) | <0.001 | **28.0** | 8.22 (5.65-11.9) | <0.001 | **7.4** | 5.33 (2.89-9.83) | <0.001 |
| Advanced stage, % <sup>B</sup> |  |  |  |  |  |  |  |  |  |
| - No | **30.6** | -- | -- | **0.6** | -- | -- | **1.3** | -- | -- |
| - Yes | **36.0** | -- | -- | **12.0** | -- | -- | **2.7** | -- | -- |
| Liver metastasis, % <sup>B</sup> |  |  |  |  |  |  |  |  |  |
| - No | **32.6** | -- | -- | **3.4** | -- | -- | 1.9 | -- | -- |
| - Yes | **39.3** | -- | -- | **30.5** | -- | -- | 3.0 | -- | -- |
| Surgical complications, % |  |  |  |  |  |  |  |  |  |
| - No | -- | -- | -- | **5.3** | 1 (ref. cat.) | -- | **0.4** | -- | -- |
| - Yes | -- | -- | -- | **7.9** | 1.24 (0.88-1.75) | 0.2 | **5.1** | -- | -- |

Abbreviations: OR: Odds Ratio; CI: Confidence Interval; ASA: American Society of Anesthesiologists. In all models, age, gender, year, region (Lombardy versus others) and cancer site (rectum versus other sites) were included a priori. <sup>A</sup> No variable other than those significant at univariate analysis was included in the final model. <sup>B</sup> Given the multicollinearity across T4 stage, advanced stage and liver metastases, three separate models were fit, each including only one of the three covariates. The model with the highest R<sup>2</sup> was kept as final. All the models were repeated with the same covariates, including region as a cluster variable, with no substantial changes in the final estimates. They were thus not shown to avoid redundancy. In all the univariate analyses, significant results (p<0.05) are indicated in bold.

The multivariate analysis including only patients who were diagnosed with liver metastasis (Table 4) confirmed that having surgery in 2020 was significantly associated with a higher risk of multiple liver metastases (OR 2.21, 95% CI 1.24-3.94, p=0.007).

**Table 4.** Multivariate analyses evaluating the potential predictors of multiple liver localisations among the patients with metastatic cancer to the liver.

| Variables | Multiple liver<br>metastases<br>(254/324)<sup>A</sup><br>% | OR (95% CI) | p |
| --- | --- | --- | --- |
| Year, % |  |  |  |
| - 2019 | 72.1 | 1 (ref. cat.) | -- |
| - 2020 | 82.2 | 2.21 (1.24-3.94) | 0.007 |
| Age class in years |  |  |  |
| - <60 | 79.8 | 1 (ref. cat.) | -- |
| - 60-69.9 | 85.4 | 1.41 (0.62-3.21) | 0.4 |
| - ≥70 | 66.0 | 0.44 (0.22-0.86) | 0.016 |
| Age, 10-year increase | -- |  |  |
| Gender |  |  |  |
| - Female | 75.1 | 1 (ref. cat.) | -- |
| - Male | 79.2 | 1.15 (0.65-2.04) | 0.6 |
| Rectum location, % |  |  |  |
| - No | 79.1 | 1 (ref. cat.) | -- |
| - Yes | 70.5 | 0.37 (0.20-0.69) | 0.002 |
| Hospital in Lombardy, % |  |  |  |
| - No | 72.9 | 1 (ref. cat.) | -- |
| - Yes | 80.8 | 1.68 (0.96-2.95) | 0.07 |
| Multiple colorectal cancers |  |  |  |
| - No | 78.3 | 1 (ref. cat.) | -- |
| - Yes | 62.5 | 0.48 (0.21-1.11) | 0.09 |
| Aggressive cancer biology, % |  |  |  |
| - No | 91.1 | 1 (ref. cat.) | -- |
| - Yes | 73.8 | 0.21 (0.07-0.57) | 0.003 |

OR: Odds Ratio. CI: Confidence Interval.
<sup>A</sup> Final model including 324 observations and adjusted for the following covariates: age, gender, year, cancer site (rectum versus other sites), Region (Lombardy versus others) - all included a priori, plus multiple colorectal cancers (yes vs no) and aggressive cancer biology (yes vs no), as they were both significant ( $p<0.05$ ) at univariate analyses.

## DISCUSSION

By March 31, 2020, Italy reported the second-highest number of confirmed COVID-19 cases (101,739, after the United States of America, 140,640 cases) and the highest number of casualties (11,591) in the world. The number of patient deaths in Italy represented almost one third (31.8%) of the total COVID-19-associated deaths worlwide.^3^ The huge impact on the healthcare system required an adequate response in terms of reallocation of resources and the imposition of a national lockdown.

Cancer screening activity was discontinued between March and May 2020, and its subsequent reactivation was neither immediate nor homogeneous across the different regions. The number of the FITs in the first 5 months of 2020 was 54.9% less than 2019.^6^ A subsequent report showed a slight improvement in the situation between June and September 2020 (overall screening program reduction: 34.2%), although at the end of September, the number of FITs carried out in Italy was still 42% lower than the previous year.^16^

This evidence increased awareness regarding the potentially detrimental effects of the screening backlogs. Worrisome predictions of increased mortality in patients diagnosed with colorectal cancer were reported in a few studies. Maringe et al. estimated an increase in the number of deaths due to colorectal cancer of between 1445 and 1563 in England, United Kingdom (UK)^12^ while Sud et al. reported that an average delay in presentation of 2 months per patient would result in 3316 to 9948 life-years lost, depending on the delay of referrals in the UK.^13^ Ricciardiello et al. showed that a screening delay beyond 6 months would be associated with an increase in more advanced stage colorectal cancers while a delay of > 12 months would result in significantly higher cancer-mortality (+12%).^11^

To date, this is the first study which has shown whether the effects of the COVID-19 outbreak resulted in worse oncologic outcomes in a real-life scenario. The present study compared the outcomes of 1755 and 1481 patients who had undergone surgery in 2019 and 2020 (March to December), respectively, in 20 referral centres located in the regions which were most severely hit during the outbreak of COVID-19 in Italy in order to discover any correlation between the first wave of SARS-CoV-2 infections, and variations in the oncologic outcomes of colorectal cancer. No evidence of an increased rate of advanced-stage cancers was shown; however, the analysis found significant discrepancies which were likely associated with the reduced screening activity and, more importantly, could have potentially affected oncologic outcomes and survival.

A higher proportion of patients undergoing surgery in 2020 were diagnosed with rectal cancer (33.8% vs. 28.3%, p=0.001). It is known that the symptoms associated with rectal cancer, such as rectal bleeding, prompt additional diagnostic tests in the population regardless of the screening programs.^17,18^ On the contrary, cancers located at more proximal sites of the colon are less evident in terms of clinical signs and are, therefore, more likely to be diagnosed at an advanced stage.^19,20^ Previous studies have showed that right-sided colon cancers are associated with worse survival, also due to their clinical subtleness.^21,22^ The higher rate of rectal cancers requiring surgery in 2020 might reflect the relative decrease in the number of patients without symptoms who would have been diagnosed using the FIT and were not, due to the discontinuation of screening. In support of this theory, the present study showed a lower rate of surgical patients with no cancer-related symptoms in 2020 (15.5% vs. 19.2%, p=0.006). The proportion of screening participants who are diagnosed with colorectal cancer who lack any symptoms reflects the capacity of the FIT to detect early-stage cancers, which translates into improved survival in these patients since screening-detected cancers are diagnosed at an earlier stage than those which are already associated with symptoms.^23–25^

A higher rate of clinical T4 stage was also found in 2020 (OR 1.38, 95% CI 1.03-1.85, p=0.029), although the oncologic relevance remains unclear since the rates of the pathological T4 stage were similar between 2019 and 2020 (4.3% vs. 4.8%, p=0.9). A clinical T4 stage was defined as the presence of cancer-induced spiculations extended over the bowel wall or the suspicion of infiltration of the surrounding structures at preoperative radiological examination. These signs do not necessarily indicate pathological cancer infiltration since they could reflect perineoplastic inflammation and fibrosis. Nevertheless, previous evidence has suggested that these radiological characteristics were significantly associated with worse survival, even in patients who were eventually diagnosed with a pathological T3 stage,^26^ implying a strong effect of the neoplastic environment on the progression and outcomes of colorectal cancer.^27^

Although the overall rate of stage IV (12.1% vs. 12.2%, p=0.9) and the specific incidence of liver metastases (10.2% vs. 10.3%, p=0.9) were similar in the two study periods (Table 1 and 2), patients who had surgery in 2020 had a significantly higher risk of being diagnosed with more than one liver metastasis, as shown in Table 4 (OR 2.21, 95% CI 1.24-3.94, p=0.007). The number of liver metastases has widely been recognised as a meaningful prognostic factor in patients affected by colorectal cancer. Afshari et al. reported a significant association between shorter survival and the number of the liver metastases from rectal cancer, and St Hill et al. showed shorter disease-free survival following hepatectomy in patients with more than one liver metastasis from colorectal cancer.^28,29^

The present study had some limitations. First, it did not represent the overall population of patients affected by colorectal cancer who had received treatment in Northern Italy during the study period since only 20 hospitals were included in the study. Second, it could have underestimated the rate of patients who were diagnosed with metastatic disease and who were referred to the oncology rather than the surgical service. Third, its retrospective nature did not allow analysing whether the COVID-19 outbreak affected the time between the onset of symptoms and the referral to surgery. Finally, the short time range of the study might have prevented the observation of significantly more advanced cancer.

However, to date, this is the only study comparing patients undergoing surgery for colorectal cancer before and during the COVID-19 outbreak in one of the countries which suffered the most in terms of infected patients and mortality during 2020. Although no gross variation in the oncologic stage was observed, the study reported worrisome evidence regarding an increased risk of advanced colorectal cancer in patients who were diagnosed in 2020 in terms of associated symptoms, cancer location, clinical T4 stage, and number of liver metastases.

In spite of the fact that SARS-CoV-2 vaccine campaigns are being carried out in many countries, no global response have been proposed, and the pandemic is far from being resolved.^30^ In particular, the risk of significant virus mutations makes the possibility of future outbreaks more than concrete.^31^ While it is still not conclusive whether the outcome variations which have been identified in the present study will impact the long-term survival of patients, it is clear that large-scale interventions are required in order to alleviate the long-term effects of the COVID-19 pandemic on the diagnostic delay of patients affected by colorectal cancer.

## Data Availability

Data will be shared upoen reasonable request by the following access criteria: with other researchers for conducting legitimate scientific research (such as meta-analyses); with investigator support, after approval of a proposal sent to the principal investigator.

## Funding

No funding was received for the study.

## Conflicts of interest

The authors have declared no conflicts of interest.

## Notes

### Competing Interest Statement

The authors have declared no competing interest.

### Clinical Trial

NCT04712292

### Funding Statement

No funding was received for the present study

### Author Declarations

Comitato Etico di Area Vasta Emilia Centro 854/2020/Oss/AOUBo 8 September 2020

