## Supplementary file 1 for "The impact of COVID-19 on the oncologic outcomes of 3236 patients undergoing ColoRectal Cancer surgery in Northern Italy in 2019 and 2020 (COVID-CRC): results of a multicentric comparative cohort study"

**Supplementary File 1. Number of cases and variation between March-December 2019 and March-December 2020 in each participating centre** and by Region

| **Centre** (see authors’ affiliation for details) | Region | Covid-treating  Centre | Elective surgery  reduction | Number of cases  (2019) | Number of cases  (2020) | Variation in cases  (%) |
| --- | --- | --- | --- | --- | --- | --- |
| ^1^ Azienda Ospedaliero Universitaria di Bologna | Emilia-Romagna | Yes | > 50% | 144 | 111 | -22.9 |
| ^5^ IRCCS Humanitas Research Hospital | Lombardy | Yes | < 50% | 86 | 86 | 0 |
| ^7^ AOU Città della Salute e della Scienza | Piedmont | Yes | < 50% | 149 | 127 | -14.8 |
| ^8^ University of Padua | Veneto | Yes | > 50% | 129 | 147 | +14 |
| ^9^ Azienda Ospedaliero-Universitaria di Bologna (Maggiore) | Emilia-Romagna | Yes | > 50% | 122 | 135 | +10.7 |
| ^10^ Fondazione Poliambulanza Hospital | Lombardy | Yes | > 50% | 139 | 102 | -26.6 |
| ^11^ San Raffaele Scientific Institute | Lombardy | Yes | > 50% | 129 | 97 | -24.8 |
| ^12^ Ospedale Mauriziano Umberto I | Piedmont | Yes | > 50% | 119 | 103 | -13.4 |
| ^13^ Policlinico San Matteo | Lombardy | Yes | > 50% | 126 | 96 | -23.8 |
| ^14^ Istituto Nazionale dei Tumori | Lombardy | No | 0% | 82 | 47 | - 42.7 |
| ^15^ Academic Hospital of Trieste | Friuli-Venezia-Giulia | Yes | < 50% | 64 | 62 | -3.1 |
| ^16^ Istituto Oncologico Veneto | Veneto | No | 0% | 60 | 64 | +6.7 |
| ^17^ Ospedale Maggiore Policlinico Milano | Lombardy | Yes | > 50% | 64 | 57 | -10.9 |
| ^18^ ASST Spedali Civili di Brescia | Lombardy | Yes | > 50% | 60 | 43 | -28.3 |
| ^19^ Humanitas Gavazzeni Bergamo | Lombardy | Yes | > 50% | 49 | 53 | +1.1 |
| ^20^ ASST Santi Paolo e Carlo | Lombardy | Yes | > 50% | 75 | 19 | -74.7 |
| ^21^ ASST Fatebenefratelli Sacco | Lombardy | Yes | > 50% | 51 | 39 | -23.5 |
| ^22^ ASST Rhodense. Ospedale di Rho | Lombardy | No | < 50% | 35 | 44 | +25.7 |
| ^23^ Policlinico San Marco | Lombardy | Yes | > 50% | 49 | 27 | - 44.9 |
| ^24^ University of Brescia | Lombardy | Yes | > 50% | 30 | 22 | -26.7 |
| **By Region** |  |  |  |  |  |  |
| Lombardy |  |  |  | 975 | 732 | -24.9 |
| Piedmont |  |  |  | 261 | 230 | -11.8 |
| Emilia-Romagna |  |  |  | 266 | 246 | -7.5 |
| Veneto and Friuli-Venezia-Giulia |  |  |  | 253 | 273 | +7.9 |
| **Total** |  |  |  | **1755** | **1481** | **-15.6** |
