## Supplementary file 2 for "The impact of COVID-19 on the oncologic outcomes of 3236 patients undergoing ColoRectal Cancer surgery in Northern Italy in 2019 and 2020 (COVID-CRC): results of a multicentric comparative cohort study"

**Supplementary File 2. COVID-CRC Collaborative group: authorship list**

**Project and writing group:** Matteo Rottoli ***(IRCCS Azienda Ospedaliero Universitaria di Bologna, Alma Mater Studiorum University of Bologna, Bologna, Italy)***, Gianluca Pellino ***(Department of Advanced Medical and Surgical Sciences, Università degli Studi della Campania "Luigi Vanvitelli", Naples, Italy)***, Antonino Spinelli ***(Department of Biomedical Sciences, Humanitas University, IRCCS Humanitas Research Hospital, Rozzano, Milan, Italy)***, Lamberto Manzoli ***(Department of Medical Sciences, University of Ferrara, Ferrara, Italy),*** Gilberto Poggioli ***(IRCCS Azienda Ospedaliero Universitaria di Bologna, Alma Mater Studiorum University of Bologna, Bologna, Italy)***

**Statistics:** Maria Elena Flacco, Lamberto Manzoli ***(Department of Medical Sciences, University of Ferrara, Ferrara, Italy)***

**Collaborators:**

Mario Morino (**PI)**, Marco Allaix, Gaspare Cannata, Erica Lombardi, Carlo Alberto Ammirati, Chiara Piceni ***(AOU Città della Salute e della Scienza, Turin, Italy)***; Salvatore Pucciarelli (**PI**), Francesco Marchegiani, Gaya Spolverato, Giacomo Ghio, Gaia Zagolin, Andrei Dorin Dragu ***(First Surgical Clinic, Department of Surgical, Oncological, and Gastroenterological Sciences, University of Padua, Padua, Italy)***; Elio Jovine (**PI**), Raffaele Lombardi, Chiara Cipressi, Maria Fortuna Offi, Cristina Larotonda ***(Division of General and Emergency Surgery, IRCCS Azienda Ospedaliero-Universitaria di Bologna, Bologna, Italy)***; Matteo Rottoli (**PI**), Gilberto Poggioli, Dajana Cuicchi, Paolo Bernante, Angela Romano, Marta Tanzanu, Angela Belvedere, Daniele Parlanti, Anna Paola Pezzuto, Gabriele Vago, Antonio Lanci Lanci, Iris Shari Russo, Tommaso Violante, Ludovica Maurino ***(IRCCS Azienda Ospedaliero Universitaria di Bologna, Alma Mater Studiorum University of Bologna, Bologna, Italy)***; Moh’d Abu Hilal (**PI**), Augusto Barbosa, Carlo Tonti, Roberta La Mendola ***(Fondazione Poliambulanza Hospital, Brescia, Italy)***; Riccardo Rosati (**PI**), Ugo Elmore, Lorenzo Gozzini, Andrea Cossu, Mattia Molteni, Paolo Parise, Francesco Puccetti ***(Department of Gastrointestinal Surgery, IRCCS San Raffaele Scientific Institute and San Raffaele Vita-Salute University, Milan, Italy)***; Alessandro Ferrero (**PI**), Michela Mineccia, Marco Palisi, Federica Gonella, Francesco Danese ***(Ospedale Mauriziano Umberto I, Turin, Italy)***; Andrea Pietrabissa (**PI**), Tommaso Dominioni, Luigi Pugliese, Andrea Peri, Marta Botti, Benedetta Sargenti ***(Department of Surgery, University of Pavia and Fondazione IRCCS Policlinico San Matteo, Pavia, Italy);*** Antonino Spinelli (**PI**), Michele Carvello, Caterina Foppa, Elisabetta Coppola, Matteo Sacchi, Francesco Carrano ***(Department of Biomedical Sciences, Humanitas University, IRCCS Humanitas Research Hospital, Rozzano, Italy)***; Marcello Guaglio (**PI**), Maurizio Cosimelli, Luca Sorrentino, Gaia Colletti, Roberto Santalucia ***(Department of Surgery, Colorectal Surgery Unit, Fondazione IRCCS Istituto Nazionale dei Tumori, Milan, Italy)***; Nicolò de Manzini (**PI**), Paola Germani, Edoardo Osenda, Hussein Abdallah, Sara Cortinovis ***(Surgical Clinic Unit, University Hospital of Trieste, Trieste, Italy)***; Pierluigi Pilati (**PI**), Boris Franzato, Ottavia De Simoni, Genny Mattara **(*UOC Chirurgia Oncologica Esofago e vie digestive, Istituto Oncologico Veneto [IOV-IRCCS], Padua, Italy***); Elisa Cassinotti (**PI**), Luigi Boni, Ludovica Baldari, Cristina Bertani ***(Fondazione IRCCS Ca' Granda Ospedale Maggiore Policlinico Milano - Università degli Studi di Milano, Milan, Italy)***; Giusto Pignata (**PI**), Rossella D’Alessio, Jacopo Andreuccetti, Ilaria Canfora, Elisa Arici, Michele De Capua ***(ASST Spedali Civili di Brescia, Brescia, Italy)***; Orlando Goletti (**PI**), Mattia Molteni, Giorgio Quartierini, Alberto Assisi ***(Chirurgia Generale Humanitas Gavazzeni Bergamo, Italy)***; Giordano Beretta **(*Oncologia Medica, Humanitas Gavazzeni Bergamo, Italy);*** Enrico Opocher (**PI**), Andrea Pisani Ceretti, Nicolò Maria Mariani ***(ASST Santi Paolo e Carlo, Dipartimento di scienze della salute - Università degli Studi di Milano, Milan, Italy)***; Piergiorgio Danelli (**PI**), Francesco Colombo, Alice Frontali, Anna Maffioli, Andrea Bondurri, Isabella Pezzoli, Alessandro Bonomi ***(ASST Fatebenefratelli Sacco, Milano; Dipartimento di Scienze Biomediche e Cliniche, Università degli Studi di Milano, Milan, Italy)***; Gianluca Sampietro (**PI**), Carlo Corbellini, Carlo Alberto Manzo, Leonardo Lorusso (***Divisione di Chirurgia Generale ed Epato-Bilio-Pancreatica. ASST Rhodense. Ospedale di Rho, Monumento ai Caduti, Rho, Milan, Italy)***; Stefano Olmi (**PI**), Matteo Uccelli, Marta Bonaldi, Giovanni Carlo Cesana ***(Chirurgia Generale ed Oncologica, Policlinico San Marco GSD, Zingonia, Bergamo, Italy)***; Nazario Portolani (**PI**), Sarah Molfino, Federico Gheza, Marie Sophie Alfano, Enrica Avezzù Pignatelli ***(Department of Clinical and Experimental Sciences, Surgical Clinic, University of Brescia, Brescia Italy)***
